# A polygenic risk score predicts mosaic loss of chromosome Y in circulating blood cells

**DOI:** 10.1101/2021.06.15.21258995

**Authors:** Moeen Riaz, Jonas Mattisson, Galina Polekhina, Andrew Bakshi, Jonatan Halvardson, Marcus Danielsson, Adam Ameur, John McNeil, Lars A. Forsberg, Paul Lacaze

## Abstract

Mosaic loss of Y chromosome (LOY) in leukocytes is associated with risk for death and disease in men. We investigated a polygenic risk score (PRS) for LOY comprising 156 previously associated germline variants, in 5131 men aged ≥70 years. Levels of LOY were estimated using microarrays and validated by whole genome sequencing. After adjusting for covariates, the PRS was a significant predictor of LOY (odds ratio 1.74). Men in the highest quintile of the PRS distribution had >5-fold higher risk of LOY than the lowest. A PRS for LOY could become a useful tool for risk prediction and targeted intervention.

## Introduction

Mosaic loss of chromosome Y (LOY) refers to acquired Y-aneuploidy in a fraction of somatic cells. Population studies have identified LOY as the most common somatic change that occurs in circulating white blood cells of older men^1-10^. In serially studied men, the fraction of blood cells with LOY typically increases in frequency over time^2,8-10^. For example, at least 40% of 70 year old men in the UK Biobank (UKBB) were affected by LOY at baseline^5^. Single-cell analyses have identified that leukocytes with LOY are found in every studied older subject^11^. Epidemiological investigations show that the presence of LOY in blood leukocytes is associated with increased risk for all-cause mortality^2,12^ and a range of common diseases in men, such as hematological and non-hematological cancer^2,10,13-17^, Alzheimer’s disease^3^, autoimmune diseases^18,19^, cardiovascular events^12,20^, age-related macular degeneration^21^ and type 2 diabetes^12^. Recent work and the diverse range of associated outcomes suggest that LOY could act as a biomarker of genomic instability^4,5^ as well as be linked with direct physiological effects; through impaired functions of affected leukocytes^2-6,11,17,22-24^. Hence, identification of men with LOY occurring in peripheral blood could help to pinpoint men in the general population who are at the highest risk of common disease from an earlier age, for targeted intervention.

In addition to age, LOY is associated with smoking and air pollution, as well as other lifestyle factors^4,9,12,25-27^. Furthermore, recent genome-wide association studies (GWAS) have identified up to 156 independent germline variants associated with risk of LOY occurring in leukocytes^4-6,25,27^. The LOY-associated germline risk variants are primarily enriched in genes related to DNA damage, cell-cycle regulation and cancer susceptibility^4,5^. These variants can now be used to calculate a polygenic risk score (PRS) to predict individual propensity to be affected with LOY and thus, add genetic predisposition as a measurable risk factor for LOY beyond age and environmental exposures. The objective of this study was to calculate a novel PRS for LOY using previously the established germline risk variants (Table S1) and to assess the predictive performance of this score in a large independent population of men aged 70 years and older. Our hypothesis was that a PRS for LOY could be used to improve risk prediction for LOY as men age, which in turn may help identify men with increased vulnerability for chronic and common disease, who could benefit from earlier targeted interventions in the future.

## Results

### Baseline Characteristics

The characteristics of the sample population are presented in Table 1. A total of 5131 DNA samples from males aged 70 years and older passed all QC metrics and were available for LOY analysis. The threshold for scoring of individuals with LOY was an mLRRY value based on array intensity data below -0.06, representing LOY in at least 8.6% of the studied blood cells in a sample. Current smokers constituted a small percentage of the population (3.5%) and the majority of participants were current alcohol users (85.3%). The frequency of LOY among all participants was 27.2% based on the binary LOY threshold and we observed higher prevalence of LOY with age; affecting more than half of the participants aged 85 or older (Table S2, Figures S1 and S2). Among the baseline characteristics, we found significant differences between men with and without LOY for age, smoking and alcohol use using the binary threshold (Table 1). No evidence of association between LOY and randomization to aspirin treatment was found.

**Table 1.** Characteristics of the Sample Population.

|  | All<br>(n=5131) | LOY No<br>(n=3739) | LOY Yes<br>(n=1392) | *p*-value* |
| --- | --- | --- | --- | --- |
| **Age at Enrolment** |  |  |  |  |
| Mean (SD) | 74.9 (4.18) | 74.4 (3.84) | 76.3 (4.73) | <0.001 |
| **Smoking** |  |  |  |  |
| Never | 2253 (43.9%) | 1680 (44.9%) | 573 (41.2%) | <0.001 |
| Former | 2699 (52.6%) | 1952 (52.2%) | 747 (53.7%) |  |
| Current | 179 (3.5%) | 107 (2.9%) | 72 (5.2%) |  |
| **Alcohol** |  |  |  |  |
| Never | 461 (9.0%) | 359 (9.6%) | 102 (7.3%) | 0.04 |
| Former | 294 (5.7%) | 211 (5.6%) | 83 (6.0%) |  |
| Current | 4376 (85.3%) | 3169 (84.8%) | 1207 (86.7%) |  |
| **Treatment** |  |  |  |  |
| Placebo | 2564 (50.0%) | 1878 (50.2%) | 686 (49.3%) | 0.57 |
| Aspirin | 2567 (50.0%) | 1861 (49.8%) | 706 (50.7%) |  |
| **BMI (kg/m2)** |  |  |  |  |
| Normal | 1063 (21.0%) | 754 (20.5%) | 309 (22.5%) | 0.07 |
| Underweight | 38 (0.8%) | 23 (0.6%) | 15 (1.1%) |  |
| Overweight | 2633 (52.1%) | 1921 (52.2%) | 712 (51.9%) |  |
| Obese | 1298 (25.7%) | 969 (26.3%) | 329 (24.0%) |  |
| Missing | 18 (0.4%) | 12 (0.3%) | 6 (0.4%) |  |
| * Using the LOY binary variable, t-test or chi-square test we performed for continuous and categorical variables, respectively. |  |  |  |  |

### Comparison of PRS distribution in men with and without LOY

We first sought to determine whether the overall PRS distribution in men with LOY had shifted compared to men without LOY. To investigate this, we plotted the PRS distributions side-by-side as density plots (Figure 1) and tested for differences in the mean PRS distribution between the two groups, adjusted for age, smoking and alcohol use. We found that men with LOY displayed on average a higher PRS, as the mean distribution in men with LOY was shifted rightwards, versus men without LOY (ANCOVA, *p*<0.001).

**Figure 1.**
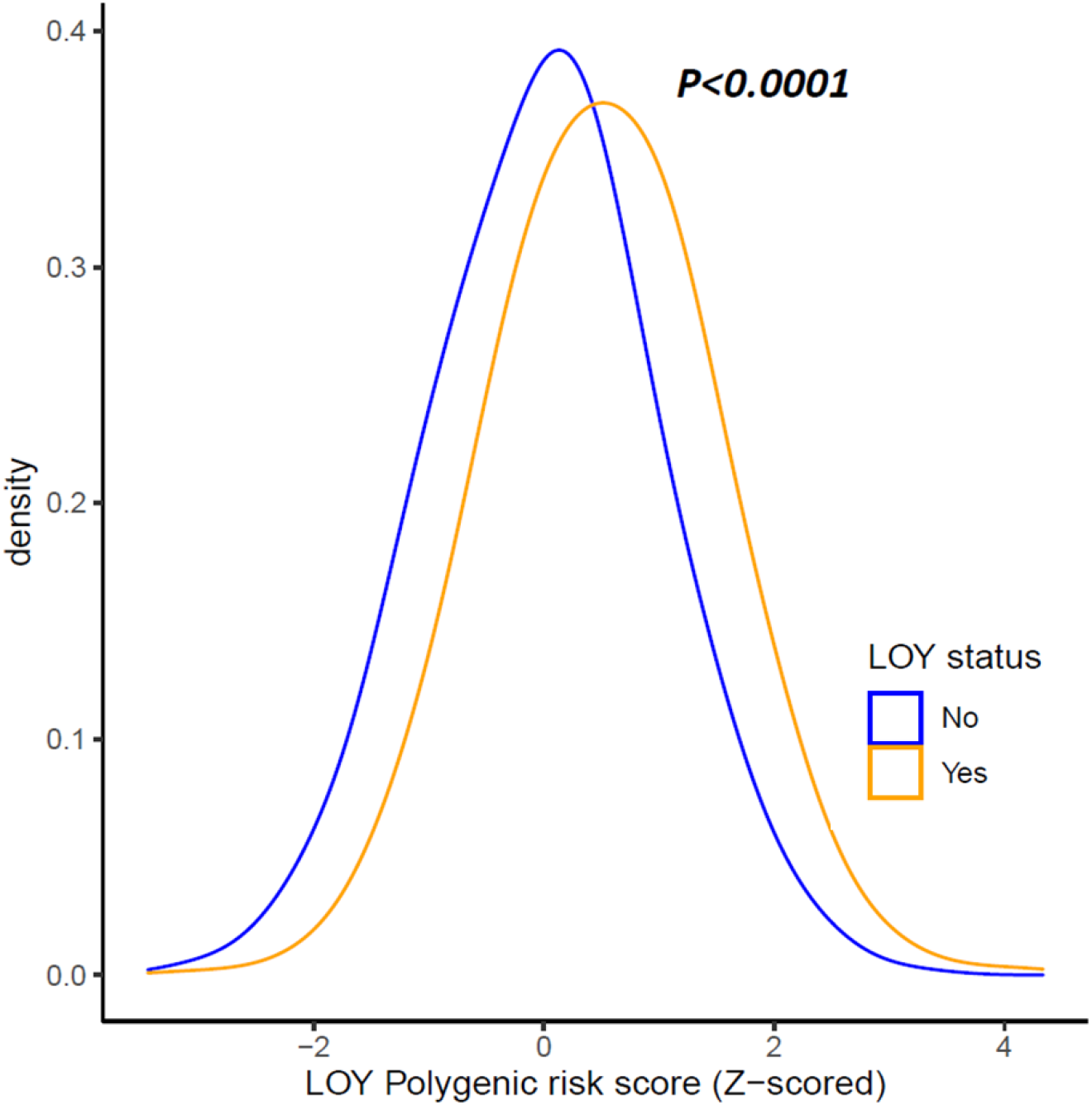
The distributions of polygenic risk scores for LOY (LOY-PRS) visualized by density plots among men with and without LOY. The *p*-value was calculated for the mean difference between the PRS distribution for participants with LOY (orange) and without LOY (blue) using ANCOVA, adjusted for age, smoking and alcohol use.

### Association of a Polygenic Risk Score with LOY mosaicism

Next, we tested for association between the LOY-PRS as a continuous variable and the binary LOY score. For each standard deviation increase in the PRS, we observed an odds ratio (OR) of 1.74 higher risk of LOY (95% confidence intervals [CI]=1.62-1.86, *p*<0.001) after adjustment for age, smoking and alcohol use (Table 2). After this, we explored the LOY-PRS as a predictor of LOY risk in models adjusted for confounding effects of age, smoking and alcohol use. First, we investigated the predictive power of each risk factor independently, by comparing the area under the curve (AUC) in the separate models, in which LOY-PRS displayed the largest AUC (Table S3). Then we compared the AUC of two LOY prediction models combining different risk factors; one including only age, smoking and alcohol use (AUC=0.63, CI=0.61-0.65) and the second including also the LOY-PRS (AUC=0.70, CI=0.68-0.71). Of note, a statistically significant improvement of the AUC was achieved by adding the LOY-PRS to the LOY risk prediction model (Figure S3, *p*<0.001).

**Table 2:** Association of a polygenic risk score for LOY predisposition (LOY-PRS) as a continuous variable, with LOY measured in 5131 men.

|  | **OR (95% CI)\*** | ***p*-value\*** |
| --- | --- | --- |
| **LOY-PRS** | 1.74 (1.62; 1.86) | **<0.0001** |
| **Age, (years)** | 1.11 (1.09; 1.13) | **<0.0001** |
| **Smoking** |  |  |
| Never/Former | Reference |  |
| Current | 2.13 (1.53; 2.94) | **<0.0001** |
| **Alcohol** |  |  |
| Never/Former | Reference |  |
| Current | 1.21 (1.01; 1.47) | **0.03** |

\* OR: odds ratios and *p*-values assessed using logistic regression.

We then analysed the LOY-PRS as a categorical variable, comparing risk of LOY for participants in the lowest quintile of the PRS distribution (Q1, reference) versus those in the highest quintile of the distribution (Q5, high-risk group) and the middle 21-80% (Q2-4, middle group). We found that men in highest quintile of the PRS distribution had over 5-fold higher risk of LOY than those in the lowest (OR=5.05, CI=4.05-6.32, *p*<0.001, Table 3). Similarly, compared with the lowest quintile, men in the middle 21-80% of the PRS distribution (middle group) also had a higher risk of LOY (OR=2.23, CI=1.83-2.73, *p*<0.001, Table 3), after adjusting for age, smoking and alcohol use. The increased risk of LOY observed for men in the high and middle PRS groups, compared with the low PRS group, was similarly observed when modelling LOY as a continuous variable (Table S4).

**Table 3:** Association of a polygenic risk score for LOY predisposition (LOY-PRS) as a categorical variable (low, middle, high), with LOY measured in 5131 men.

|  | **OR (95% CI)\*** | ***p*-value\*** |
| --- | --- | --- |
| **LOY low risk PRS** | Reference |  |
| **LOY middle risk PRS** | 2.23 (1.83-2.73) | **<0.0001** |
| **LOY high risk PRS** | 5.05 (4.05; 6.32) | **<0.0001** |
| **Age, (years)** | 1.12 (1.09; 1.12) | **<0.0001** |
| **Smoking** |  |  |
| Never/Former | Reference |  |
| Current | 2.00 (1.44; 2.76) | **<0.0001** |
| **Alcohol** |  |  |
| Never/Former | Reference |  |
| Current | 1.2 (1.00; 1.45) | **0.05** |

\* OR: odds ratios and *p*-values assessed using logistic regression. The LOY-PRS group define as: low <20% , Middle 30-60% and high >80% PRS.

### Sub-group analysis by age

To further investigate whether the PRS continued to be associated with higher risk of LOY as men age (e.g. independently of age), we stratified the cohort into three age-ranges; 70-74 years, 75-79 years and 80+ years and examined the effect of the PRS in each age group separately. These analyses showed that the association between the PRS and risk of LOY remained significant in each age range, and interestingly; that the strength of the PRS prediction increased with age (Figure 2). Specifically, among participants aged 70-74 years, we observed an increased risk of LOY in the high PRS group (OR=2.35, CI=1.97-2.81, *p*<0.001) and in the middle group (OR=1.30 CI=1.13-1.50, *p*<0.001), versus the low group, after adjusting for smoking and alcohol use. Moreover, for men aged 75-79 years, we observed a stronger PRS effect than in the younger group, with a higher risk of LOY in the high PRS group (OR=4.00, CI=2.90-5.52, *p*<0.001) as well as the middle group (OR=1.55, CI=1.19-2.02, *p*<0.001). In the 80+ age-range, despite smaller participant numbers, we observed similar odds ratios compared with the 75-79 age-range, with higher risk of LOY in the high PRS group (OR=4.14, CI=2.12-8.08, *p*<0.001) and the middle group (OR=2.09, CI=1.19-3.67, *p*<0.010).

**Figure 2:**
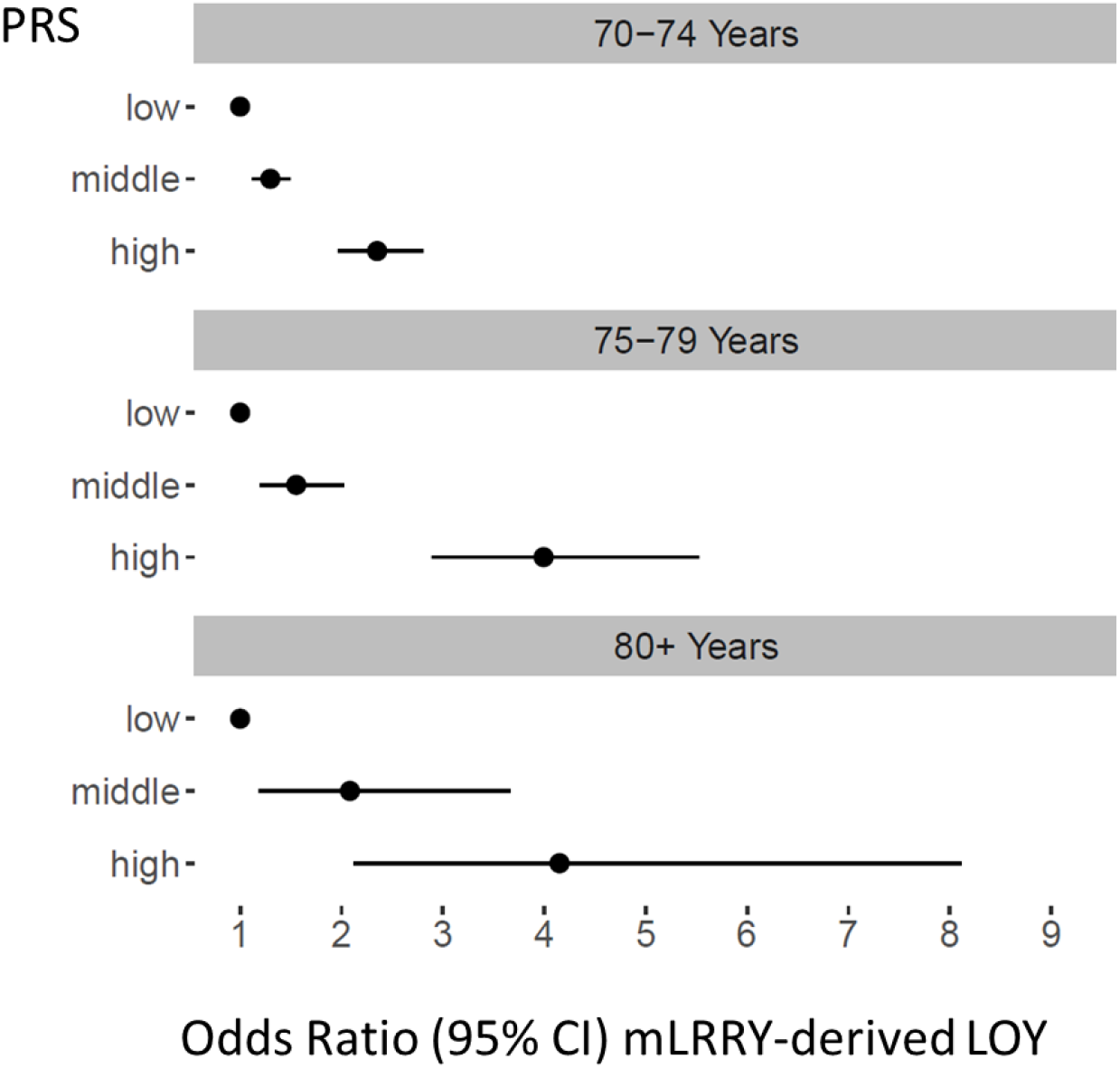
The predictive power of the LOY-PRS increase with the age. The age dependence was evaluated by comparing results derived from the age groups 70-74, 75-79 and 80+ years, respectively. Within each age group, the predictive power of the PRS (estimated with odds ratios) is shown for men with low PRS (Q1 of PRS distribution; i.e. 0-20%), middle PRS (Q2-4; 21-80%) and high PRS (Q5; 81-100%).

### Validation of LOY using whole genome sequencing data

The SNP array derived LOY estimation was validated using an orthogonal genomic technology. We performed a concordance analysis of LOY calls detected by microarray versus LOY calls based on whole genome sequencing (WGS) read depth, for a sub-set of 947 men for whom WGS data was available. The microarray-derived and WGS-derived LOY calls were highly correlated (Pearson correlation coefficient = 0.98) (Figure S4).

## Discussion

Recent studies have provided insights into the potential disease mechanisms that could help explain why men affected with LOY in blood cells live shorter lives. First, GWAS has identified germline variants associated with risk of LOY in leukocytes. Many of these risk variants are shared with loci for various diseases, and highlight genes involved in cell cycle regulation, DNA damage response and cancer susceptibility^4-6,25,27^. This ‘common soil’ of genetic predisposition helps, at least in part, to explain why men with LOY in peripheral blood display an increased risk for disease, that may be mediated by a age-related genomic imbalance in somatic tissues^5^. Second, it has been proposed that LOY in leukocytes could be linked with risk for disease in other organs by impaired immune functions of affected leukocytes^2,3,5,7,9,22,23,28^. This hypothesis is supported by studies suggesting involvement of chromosome Y in processes such as leukocyte development and function as well as transcriptional regulation^6,11,28-34^. For example, patients diagnosed with prostate cancer and Alzheimer’s disease have been shown to be affected with LOY in different types of immune cells, indicating a disease-specific link^11^. Furthermore, observations of extreme down-regulation of chromosome Y genes (EDY) in different types of cancers^35^ and in Alzheimer’s disease^36^ demonstrates that expression of Y-linked genes could be important in the context of disease protection. Moreover, almost 500 autosomal genes have been shown to display LOY-associated transcriptional effect (LATE) by dysregulation in peripheral leukocytes with LOY, including many genes important for physiological immune functions^11^. In aggregate, LOY in blood cells could either act as a barometer of genomic imbalance in- and outside of the hematopoietic system and furthermore, it is plausible that immune cells with this aneuploidy could be directly linked with disease etiology in human disease conditions with an immunological component.

In this study, we examined the predictive performance of a polygenic risk score (PRS) based on 156 previously-associated risk variants for LOY^5^ using array data from 5131 healthy men aged 70 years and older. We found that the PRS was a significant predictor of LOY after adjusting for confounders, such as age, smoking and alcohol use. For each standard deviation increase in the PRS, we observed a 1.7-fold higher risk of LOY. Men in the highest quintile of the PRS distribution had, on average, more than 5-fold higher risk of LOY compared with men in the lowest quintile of the distribution. A risk prediction model for LOY was improved significantly by the addition of the PRS to conventional risk factors such as age, smoking and alcohol use. Thus, regardless of potential mechanisms behind associations between LOY in blood cells and various outcomes discussed above, the results presented here show that the germline variation captured by the PRS can help identify men at highest risk of LOY in leukocytes. These results have implications for improved risk stratification and targeted intervention in ageing men.

We defined LOY using a microarray-derived signal intensity threshold, which corresponded to >8.6% of cells losing the Y chromosome. We validated the microarray-derived LOY calls using WGS data. Based on the threshold, we found that the prevalence of LOY in the overall study population was 27.2%. After stratification by age, the frequency of men with LOY was 21%, 32%, 44% and 51% in men aged 70-74, 75-79, 80-84 and 85 years or older, respectively, consistent with previous reports^1-10^. Stratified analysis performed within age groups showed that the PRS was a significant predictor of LOY across all ages, with stronger predictive power in older men. This result fits well with previous data showing an accumulation of LOY with age, in the general population and an increased frequency of leukocytes with LOY in the blood of serially studied men^2,8-10^.

Strengths of our study include the well-characterized, older study population (mean age of 75 years at enrolment) with genotyping and WGS data available. A further strength is the ability of the ASPREE cohort to act as an independent validation of a PRS derived from germline variants identified from the UK Biobank population. Limitations of our study include the potential for survivorship bias in participant ascertainment, with individuals enrolled into the ASPREE study likely being healthier and at lower risk of disease than individuals from the general population in the same age range. Further, given that the majority of ASPREE participants were individuals of European genetic descent, this may limit the generalizability of our results to other ethnicities.

In summary, this is the first study to predict risk for LOY in leukocytes based on a PRS derived from germline variants, in a large population of older men. Mosaic LOY aneuploidy in leukocytes is associated with morbidity and mortality in populations of aging men, and constitutes a promising biomarker for general disease vulnerability. We report here that the inherited genetic make-up of individuals could be used to identify high-risk men with elevated likelihood of being affected with LOY, which could benefit early diagnosis and prevention of common disease in the future. Implementation of a PRS for LOY risk prediction could promote earlier diagnoses of common disease, as well as enable risk stratification of men who would benefit more from early targeted intervention for a range of LOY-associated diseases.

## Methods

### Study population

This study was comprised of male participants of the ASPREE trial, a randomized, placebo-controlled trial investigating the effect of daily 100mg aspirin on disability-free survival in healthy older individuals^37-39^. ASPREE inclusion criteria and baseline characteristics have been reported previously^40^. Briefly, individuals over the age of 70 years were enrolled, who had no previous history or current diagnosis of atherothrombotic cardiovascular disease events, dementia, loss of independence with basic activities of daily living, or any serious illness likely to cause death within five years, as confirmed by a general practitioner assessment. ASPREE participants also passed a global cognition screen at enrolment, scoring >77 on the Modified Mini-Mental State (3MS) Examination. Participants were recruited between 2010-2014 through general (family) practitioners in Australia and trial centres in the US. Informed consent for genetic analysis was obtained, with ethical approval from the Alfred Hospital Human Research Ethics Committee (390/15) and site-specific Institutional Review Boards in the US.

### Microarray genotyping and imputation

We genotyped DNA from 6,140 peripheral blood samples provided by male participants at the time of study enrolment using the Axiom 2.0 Precision Medicine Diversity Research Array (PMDA) following standard protocols. To estimate population structure and ethnicity, we performed principal component analysis using the 1000 Genomes reference population (Figure S5)^41^. Variant-level quality control included filters on >90% genotyping rate and Hardy Weinberg-equilibrium, using plink version 1.9^42^. Genotype data was imputed using the TOPMed server^43-45^. Post-imputation QC removed any variants with low imputation quality scores (r2<0.3).

### Estimation of LOY from microarray data

The level of LOY mosaicism in each participant was estimated using microarray intensity data from male-specific chromosome Y probes (MSY) as described in Supplementary Materials and Figures S6-S8. Briefly, Log R Ratio (LRR) output can be used to quantify copy number states from microarray data. The LRR is calculated as the logged ratio of the observed probe intensity to the expected intensity and observed LRR deviation in a specific genomic region is therefore indicative of copy number change. After quality control steps based on genotyping quality, sex, relatedness and ancestry; a total of 5131 male samples were retained for LOY analysis. For each sample, we first calculated the mLRRY as the median of the LRR values of the 488 Y-specific probes on the array, i.e. located within the MSY. The mLRRY is a continuous estimate of LOY; a value close to zero indicate a normal state while samples with LOY display mLRRY values below zero. To score samples with or without LOY we defined a threshold based on technical variation as described previously^9^ and the percentage of cells with LOY in each participant was calculated^8^. We considered LOY as a continuous and categorical/binary variable in different analyses. Individuals with mLRRY less than -0.06 (equivalent to the 0.5th percentile of experimental error distribution) corresponding to >8.6% of blood cells having LOY were considered as having LOY as a categorical variable.

### Estimation of LOY from whole genome sequencing data

We used whole genome sequencing (WGS) data that was available from 2795 ASPREE participants (male and female) through the Medical Genome Reference Bank project^46,47^. WGS data was produced on the Illumina HiSeq X system with an average of 30x sequencing coverage as described previously^47^. We compared microarray-derived and WGS-derived LRR calls using Pearson correlation in 947 male participants for whom both microarray and WGS data was available. LOY estimation from the WGS data was based on read depth, rather than LRR intensity differences. WGS data was analysed using the Control-FREEC software (version 11.5)^48^ (details see Supplementary Materials).

### Calculation of polygenic risk score

The LOY polygenic risk score (LOY-PRS) was generated using 156 genome-wide significant variants previously associated with LOY^5^. A total of 123 variants passed genotyping and imputation QC thresholds and were present in the ASPREE imputed SNP array data set and were used to calculate the PRS (Table S1). Plink version 2 was used to calculate the LOY-PRS as weighted sum of log odd ratios and effect alleles for each variant^49^. We categorized the LOY-PRS distribution into three groups based on quintiles (Q); low (Q1, 0-20%), middle (Q2-4, 21-80%) and high (Q5, 81-100%) risk.

### Statistical analysis

Baseline characteristics included age, smoking (current/former and never), alcohol use, body mass index (BMI) and treatment assignment (aspirin or placebo). Using the LOY binary variable, we performed a t-test or chi-square test for baseline continuous and categorical variables, respectively. We assessed the difference in LOY distribution by age using the Wilcoxon Test. The LOY-PRS distribution was Z-score standardised to have a mean 0 (SD 1) and tested for association in men with and without mLRRY-derived LOY using ANCOVA adjusting for age smoking and alcohol use. We than performed multivariable regression model for per standard deviation increase in LOY-PRS with mLRRY-derived LOY dichotomous and linear variable adjusting for baseline characteristics. In a separate regression model, the risk of mLRRY derived LOY (binary or continuous variable) was assessed between LOY-PRS categories using quintiles (Q) of the PRS distribution, considering the low-risk PRS group (Q1, 0-20%) as a reference, comparing against middle (Q2-4, 21-80% and high (Q5, 81-100%) risk groups. For sub-group analysis the LOY-PRS risk categories were further stratified into three age groups; 70-74 years, 75-79 years and 80+ years. Finally, the area under the curve (AUC) was calculated for age, smoking and alcohol use followed by adding LOY-PRS using receiver-operating-characteristics (ROC). We used DeLong’s test to compare the two ROC curves^50^. All analysis is performed using R version 4.0.3.

## Supporting information

Supplement Details

## Data Availability

The data that support the findings of this study are available from the corresponding author upon request

## Acknowledgements

This work was supported by an ASPREE Flagship cluster grant (including the Commonwealth Scientific and Industrial Research Organization, Monash University, Menzies Research Institute, Australian National University, University of Melbourne); and grants (U01AG029824 and U19AG062682) from the National Institute on Aging and the National Cancer Institute at the National Institutes of Health, by grants (334047 and 1127060) from the National Health and Medical Research Council of Australia, and by Monash University and the Victorian Cancer Agency. PL is supported by a National Heart Foundation Future Leader Fellowship (ID 102604). JJM is supported by an Investigator grant from the National Health and Medical Research Council of Australia (1173690). This result is part of a project L.A.F has received funding from the European Research Council (ERC) under the European Union’s Horizon 2020 research and innovation programme (Grant agreements No. 679744 and 101001789). L.A.F is also supported by grants from the Swedish Research Council (#2017-03762), the Swedish Cancer Society (# 20-1004) and Kjell och Märta Beijers Stiftelse. Data handling and computations were enabled by resources provided by the Swedish National Infrastructure for Computing (SNIC) at Uppsala Multidisciplinary Center for Advanced Computational Science (UPPMAX) partially funded by the Swedish Research Council (#2018-05973), under the project SNIC (sens2019020).

## Author Contributions

MR, JM: Conceptualization, Data curation, Formal Analysis, Methodology, Investigation, Software, Writing – original draft. GP, AB, JH, MD: Data curation, Formal Analysis, Methodology. AA, JJM: Data curation, Funding acquisition, Supervision. PL, LAF: Conceptualization, Data curation, Funding acquisition, Methodology, Supervision, Writing original draft. All authors were involved in writing (review and editing).

## Competing Interests statement

L.A.F. is co-founder and shareholder in Cray Innovation AB.

## Notes

### Author Declarations

Informed consent for genetic analysis was obtained, with ethical approval from the Alfred Hospital Human Research Ethics Committee (390/15) and site-specific Institutional Review Boards in the US

## References

1. UKCCG. Loss of the Y chromosome from normal and neoplastic bone marrows. United Kingdom Cancer Cytogenetics Group (UKCCG). Genes Chromosom Cancer 5, 83–8 (1992).

2. Forsberg, L.A. et al. Mosaic loss of chromosome Y in peripheral blood is associated with shorter survival and higher risk of cancer. Nat Genet 46, 624–8 (2014).

3. Dumanski, J.P. et al. Mosaic Loss of Chromosome Y in Blood Is Associated with Alzheimer Disease. Am J Hum Genet 98, 1208–19 (2016).

4. Wright, D.J. et al. Genetic variants associated with mosaic Y chromosome loss highlight cell cycle genes and overlap with cancer susceptibility. Nat Genet 49, 674–679 (2017).

5. Thompson, D.J. et al. Genetic predisposition to mosaic Y chromosome loss in blood. Nature 575, 652–657 (2019).

6. Terao, C. et al. GWAS of mosaic loss of chromosome Y highlights genetic effects on blood cell differentiation. Nat Commun 10, 4719 (2019).

7. Forsberg, L.A. et al. Mosaic loss of chromosome Y in leukocytes matters. Nat Genet 51, 4–7 (2019).

8. Danielsson, M. et al. Longitudinal changes in the frequency of mosaic chromosome Y loss in peripheral blood cells of aging men varies profoundly between individuals. Eur J Hum Genet 28, 349–357 (2020).

9. Dumanski, J.P. et al. Smoking is associated with mosaic loss of chromosome Y. Science 347, 81–83 (2015).

10. Ouseph, M.M. et al. Genomic alterations in patients with somatic loss of the Y chromosome as the sole cytogenetic finding in bone marrow cells. Haematologica (2020).

11. Dumanski, J.P. et al. Immune cells lacking Y chromosome show dysregulation of autosomal gene expression. Cell Mol Life Sci (2021).

12. Loftfield, E. et al. Predictors of mosaic chromosome Y loss and associations with mortality in the UK Biobank. Sci Rep 8, 12316 (2018).

13. Ganster, C. et al. New data shed light on Y-loss-related pathogenesis in myelodysplastic syndromes. Genes Chromosomes Cancer 54, 717–24 (2015).

14. Noveski, P. et al. Loss of Y Chromosome in Peripheral Blood of Colorectal and Prostate Cancer Patients. PLoS One 11, e0146264 (2016).

15. Machiela, M.J. et al. Mosaic chromosome Y loss and testicular germ cell tumor risk. J Hum Genet 62, 637–640 (2017).

16. Loftfield, E. et al. Mosaic Y Loss Is Moderately Associated with Solid Tumor Risk. Cancer Res 79, 461–466 (2019).

17. Asim, A. et al. Investigation of LOY in Prostate, Pancreatic and Colorectal Cancers in males: A case-control study. Expert Rev Mol Diagn (2020).

18. Persani, L. et al. Increased loss of the Y chromosome in peripheral blood cells in male patients with autoimmune thyroiditis. J Autoimmun 38, J193–6 (2012).

19. Lleo, A. et al. Y chromosome loss in male patients with primary biliary cirrhosis. J Autoimmun 41, 87–91 (2013).

20. Haitjema, S. et al. Loss of Y Chromosome in Blood Is Associated with Major Cardiovascular Events during Follow-up in Men after Carotid Endarterectomy. Circ Cardiovasc Genet 10:e001544 (2017).

21. Grassmann, F. et al. Y chromosome mosaicism is associated with age-related macular degeneration. Eur J Hum Genet 27, 36–41 (2019).

22. Forsberg, L.A., Gisselsson, D. & Dumanski, J.P. Mosaicism in health and disease - clones picking up speed. Nat Rev Genet 18, 128–142 (2017).

23. Forsberg, L.A. Loss of chromosome Y (LOY) in blood cells is associated with increased risk for disease and mortality in aging men. Human Genetics 136, 657–663 (2017).

24. Guo, X. et al. Mosaic loss of human Y chromosome: what, how and why. Hum Genet 139, 421–446 (2020).

25. Zhou, W. et al. Mosaic loss of chromosome Y is associated with common variation near TCL1A. Nat Genet 48, 563–8 (2016).

26. Wong, J.Y.Y. et al. Outdoor air pollution and mosaic loss of chromosome Y in older men from the Cardiovascular Health Study. Environ Int 116, 239–247 (2018).

27. Liu, Y. et al. Polycyclic aromatic hydrocarbons exposure and their joint effects with age, smoking, and TCL1A variants on mosaic loss of chromosome Y among coke-oven workers. Environ Pollut, Published online, 10.1016/j.envpol.2019.113655,(2019).

28. Case, L.K. et al. The Y chromosome as a regulatory element shaping immune cell transcriptomes and susceptibility to autoimmune disease. Genome Res 23, 1474–85 (2013).

29. Sun, S.L. et al. Y chromosome-linked B and NK cell deficiency in mice. J Immunol 190, 6209–20 (2013).

30. Wesley, J.D., Tessmer, M.S., Paget, C., Trottein, F. & Brossay, L. A Y chromosome-linked factor impairs NK T development. J Immunol 179, 3480–7 (2007).

31. Case, L.K. et al. Chromosome y regulates survival following murine coxsackievirus b3 infection. G3 (Bethesda) 2, 115–21 (2012).

32. Lin, S.H. et al. Mosaic chromosome Y loss is associated with alterations in blood cell counts in UK Biobank men. Sci Rep 10, 3655 (2020).

33. Maan, A.A. et al. The Y chromosome: a blueprint for men’s healthã Eur J Hum Genet 25, 1181–1188 (2017).

34. Bellott, D.W. et al. Mammalian Y chromosomes retain widely expressed dosage-sensitive regulators. Nature 508, 494–9 (2014).

35. Caceres, A., Jene, A., Esko, T., Perez-Jurado, L.A. & Gonzalez, J.R. Extreme Downregulation of Chromosome Y and Cancer Risk in Men. J Natl Cancer Inst 112, 913–920 (2020).

36. Caceres, A., Jene, A., Esko, T., Perez-Jurado, L.A. & Gonzalez, J.R. Extreme downregulation of chromosome Y and Alzheimer’s disease in men. Neurobiol Aging 90, 150 e1–150 e4 (2020).

37. McNeil, J.J. et al. Effect of Aspirin on Cardiovascular Events and Bleeding in the Healthy Elderly. N Engl J Med 379, 1509–1518 (2018).

38. McNeil, J.J. et al. Effect of Aspirin on Disability-free Survival in the Healthy Elderly. N Engl J Med 379, 1499–1508 (2018).

39. McNeil, J.J. et al. Effect of Aspirin on All-Cause Mortality in the Healthy Elderly. N Engl J Med 379, 1519–1528 (2018).

40. McNeil, J.J. et al. Baseline characteristics of participants in the ASPREE (ASPirin in Reducing Events in the Elderly) study. The Journals of Gerontology: Series A 72, 1586–1593 (2017).

41. Genomes Project, C. et al. A global reference for human genetic variation. Nature 526, 68–74 (2015).

42. Purcell, S. et al. PLINK: a tool set for whole-genome association and population-based linkage analyses. Am J Hum Genet 81, 559–75 (2007).

43. Fuchsberger, C., Abecasis, G.R. & Hinds, D.A. minimac2: faster genotype imputation. Bioinformatics 31, 782–4 (2015).

44. Taliun, D. et al. Sequencing of 53,831 diverse genomes from the NHLBI TOPMed Program. bioRxiv, 563866 (2019).

45. Das, S. et al. Next-generation genotype imputation service and methods. Nat Genet 48, 1284–1287 (2016).

46. Lacaze, P. et al. Medically actionable pathogenic variants in a population of 13,131 healthy elderly individuals. Genetics in Medicine, 1–4 (2020).

47. Pinese, M. et al. The Medical Genome Reference Bank contains whole genome and phenotype data of 2570 healthy elderly. Nature communications 11, 1–14 (2020).

48. Boeva, V. et al. Control-FREEC: a tool for assessing copy number and allelic content using next-generation sequencing data. Bioinformatics 28, 423–425 (2012).

49. Chang, C.C. et al. Second-generation PLINK: rising to the challenge of larger and richer datasets. Gigascience 4, 7 (2015).

50. DeLong, E.R., DeLong, D.M. & Clarke-Pearson, D.L. Comparing the areas under two or more correlated receiver operating characteristic curves: a nonparametric approach. Biometrics 44, 837–45 (1988).

