## Supplement Details for "A polygenic risk score predicts mosaic loss of chromosome Y in circulating blood cells"

**Riaz et al.**

| **Items** | **Page(s)** |
|---|---|
| Supplementary Methods | 2-5 |
| Figures S1-S8 | 6-13 |
| Tables S1-S6 | 14-17 |
| Supplementary references | 18 |

### Supplementary Methods

#### LOY estimation from SNP-array data

Whole blood DNA samples collected from 6,140 male ASPREE participants were genotyped using the Axiom 2.0 Precision Medicine Diversity Research Array (PMDA) following standard protocols (ThermoFisher). We followed best practice genotyping and quality control (QC) protocols from Thermo Fisher, starting from raw intensity CEL files, we used a command line custom script designed for the the Axiom PMDA array, mapped to human genome reference GRCh38, to produce variant call files.

We performed sample level QC using plink version 1.9, excluding samples for gender discordance (80 samples mismatched and excluded) using plink default F statistics threshold (≤ 0.2 female and ≥ 0.8 male), relatedness (124 indviduals excluded) using default PI-HAT threshold >0.025 to exclude one sample from each related pair.

To estimate population structure in the ASPREE cohort we performed principal component analysis (PCA) using The 1000 Genomes Project as a reference population[1]. Directly genotyped data from ASPREE and The 1000 Genomes Project 1K phase 3 (liftover to GRCh38) were merged and LD pruned ($r^2 < 0.1$) using plink version 1.9[2] followed by R package SNPrelate[3]. We calculated the Z score for first 2 principal component eigenvectors and excluded samples with ± 2SD (standard deviation) of Z score compared to their respective five reference superpopulation groups from the 1000 Genomes Project that included: Europenas, South Asians, East Asians, African American (African super population) and Hispanics (Ad Mixed American) (Figure S5). The final dataset of 12,815 samples (6,140 males) from Caucasians (Non-Finish Europeans) were selected from the ASPREE cohort for further analysis.

From the Axiom genotyping dataset we generated Log R Ratio (LRR) calls from the allele specific signal intensity data for each marker using a custom pipeline from the CEL files. Following this, the genomewide experimental quality for each sample was assessed and 340 samples were excluded based on published criteria for sample inclusion[4]. We also exclude 669 saliva samples from the data set. After the quality controls based on sex, relatedness, ancestry, blood/saliva sample and genotyping quality; a total of 5,131 male samples were retained for LOY analysis.

Following this, the level of LOY mosaicism in the 5,131 samples passing strict QC was estimated by calculation of the mLRRY, as described previously[5]. First, the mLRRY was calculated for each sample as the median of the Log R Ratio (LRR) values of the 488 Axiom probes located within the male specific part of chromosome Y (MSY, chrY:2.787.139-22.318.450; GRCh38/hg38). We also calculated an mLRRX value (median LRR of probes positioned in the non-PAR part of chromosome X) for each sample, and observed a technical covariation with mLRRY. To adjust for this potential bias for the LOY assessment, an adjustment of mLRRY values was performed by a new approach, based on the coefficients of a linear model estimated between mLRRY and mLRRX in the unadjusted dataset (Figure S6). Following this, the mLRRY values over the entire dataset was adjusted using a constant defined as the peak of the unadjusted mLRRY distribution. This operation shifts the peak of the mLRRY distribution so that samples without LOY align around zero, thus improving comparability between datasets. The constant applied in the present dataset (i.e. 0.0006827) was calculated as the peak of the local regression median of the unadjusted mLRRY distribution using kernel estimation in the *density* function in R and the smoothing bandwidth method “SJ”. We next defined a threshold in the adjusted mLRRY distribution that could be used for scoring samples with or without LOY. Samples with adjusted mLRRY values lower than the 99% confidence limit in a simulated distribution of technical mLRRY variation was scored with LOY (Figure S7). As a final step to improve interpretability of our results, we used a modified version of a published formula[6] to translate individual mLRRY values into percentage of cells with LOY in every sample, as described in Figure S8. Finally, the concordance between LOY estimation from SNP-array and whole genome sequencing data was evaluated and shown in Figure S4. The distribution of mosaic LOY in male blood samples observed in ASPREE study in relation to age is shown in Figures S1 and S2.

**Whole genome Sequecning and LOY estimation**

We performed Whole Genome Sequencing (WGS) of 2,795 ASPREE participants[7,8]. Samples were WGS on the Illumina HiSeq X system with an average of 30x sequencing coverage as described previously[8]. The sequence alignment and processing were performed using reference 1000 Genomes Phase 3 decoyed version of build 37 of the human genome through Genome analysis tool kit (GATK) pipeline[9]. The joint genotype calling was performed in a single batch using GTAK GenotypeGVCFs function. To estimate the WGS level of LOY, we used ASPREE WGS samples that passed MGRB quality control criteria that includes call rate > 98%, depth standard deviation < 10, VAF standard deviation at loci called heterozygous <1, hetero:homo variant ratio of <2, inbreed coefficient form X chromosome values between < 0.2 or > 0.8 and Singleton rate < 0.001[8]. Total 947 males from ASPREE WGS cohort were also genotyped on AXIOM SNP array chip and used to check the LOY concordance across two platforms.

To estimate LOY using WGS data, first, the program Control-FREEC (version 11.5)[10] was used to calculate a read depth ratio in 50.000 bp genomic windows on the UPPMAX Bianca cluster. The default parameters was used for other Control-FREEC settings. A mappability gem file for hg19 (read length 100 bp and up to 2 mismatches) was used in combination with the ASPREE-reference genome. To get a per chromosome read depth ratio, the median was calculated for all windows on each chromosome. Next, samples with deviating chromosomalratios was removed, i.e. suspected female samples (median X-ratio > 0.7), samples with suspected XYY aberration (median Y-ratio > 0.75) as well as all samples showing different types of abnormal autosomal read depth ratios (read depth standard deviation 1.5 IQR outside the third quartile).

LOY calling in the remaining 1137 male samples was performed from the ControlFREEC output. First, the *density* function in R with bandwidth set to "SJ", was used to find the highest density Y-ratio point (using all Y chromosome window rations) and the Scales package was used to linearly rescale the Y-ratios based on the density value. This resulted in a first estimate of LOY percentage in each sample between 0 and 100% cells with Y loss aneuploidy. To further improve these estimates, we identified normal samples (without LOY) as those showing a level of LOY within 2 standard deviations in the distribution. In these non-LOY samples, we noticed some highly variable Y-regions that were considered less informative for LOY calling. Specifically, 50.000 bp windows on chromosome Y with a standard deviation larger than 1.5 IQR from third quartile was considered highly variable and excluded. After this, the median Y-ratio was re-calculated for all male samples using only the Y-regions windows that passed this additional QC step. Thus, a final estimate of LOY percentage was achieved using the *density* function in R (bandwidth="SJ") and rescaled as described above in the first step. Finally, a binary threshold used for LOY scoring of samples was calculated following the same principle as described for the SNP array data. Finally, the concordance between LOY estimation from whole genome sequencing and SNP-array data was evaluated and shown in Figure S4.

**Figure S1.**

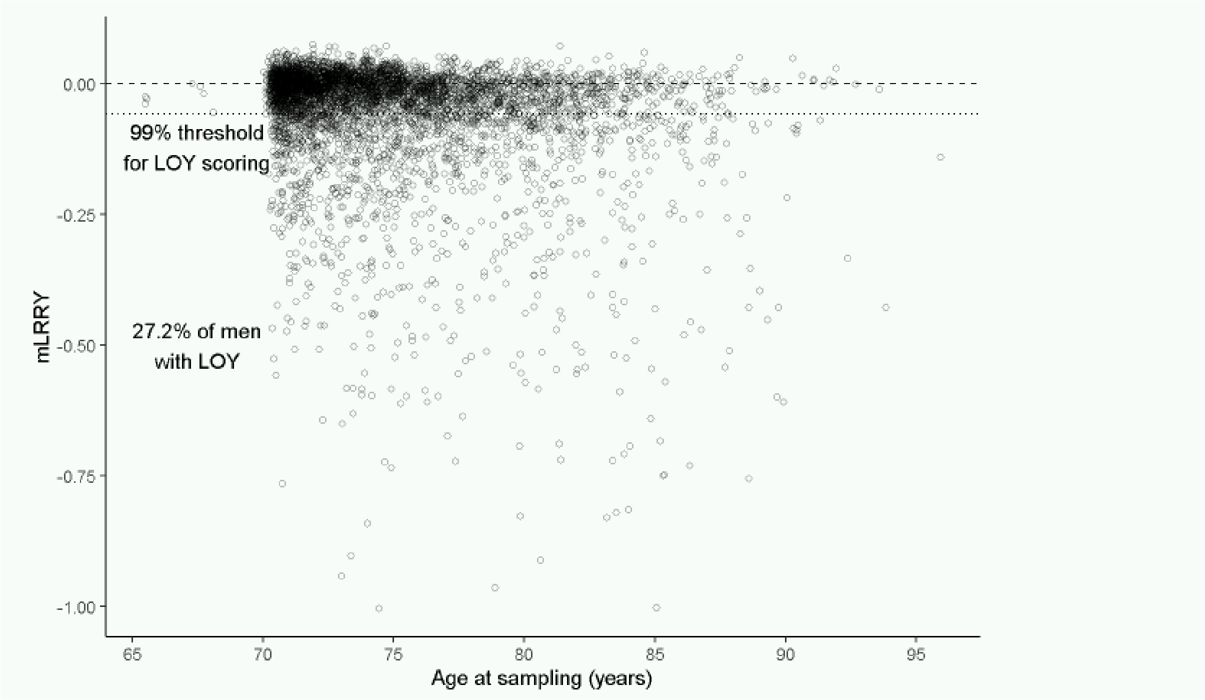

**Figure S1.** Observed level of LOY mosaicism in 5.131 ASPREE men, estimated from SNP array data and plotted in relation to age at blood sampling. The dotted line represents the threshold in the mLRRY distribution at the 99$^{th}$ percentile of experimental noise (mLRRY = -0.06) that was used for the scoring of individuals with or without LOY.

**Figure S2.**

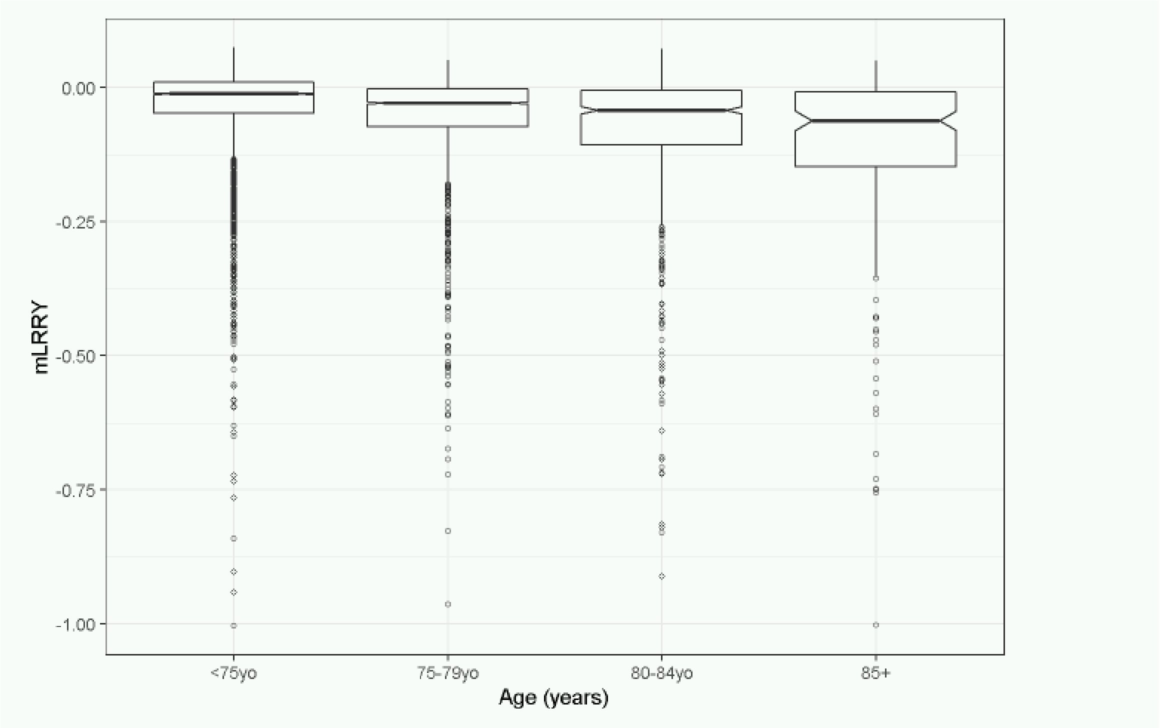

**Figure S2.** The frequency of mosaic LOY in blood samples of 5.131 ASPREE male participants in relation to age. The box and Whisker plot was generated in four age groups, the middles line in the box show the median around the interquartile ranges and top and bottom vertical lines are 95% confidence intervals.

**Figure S3.**

a)

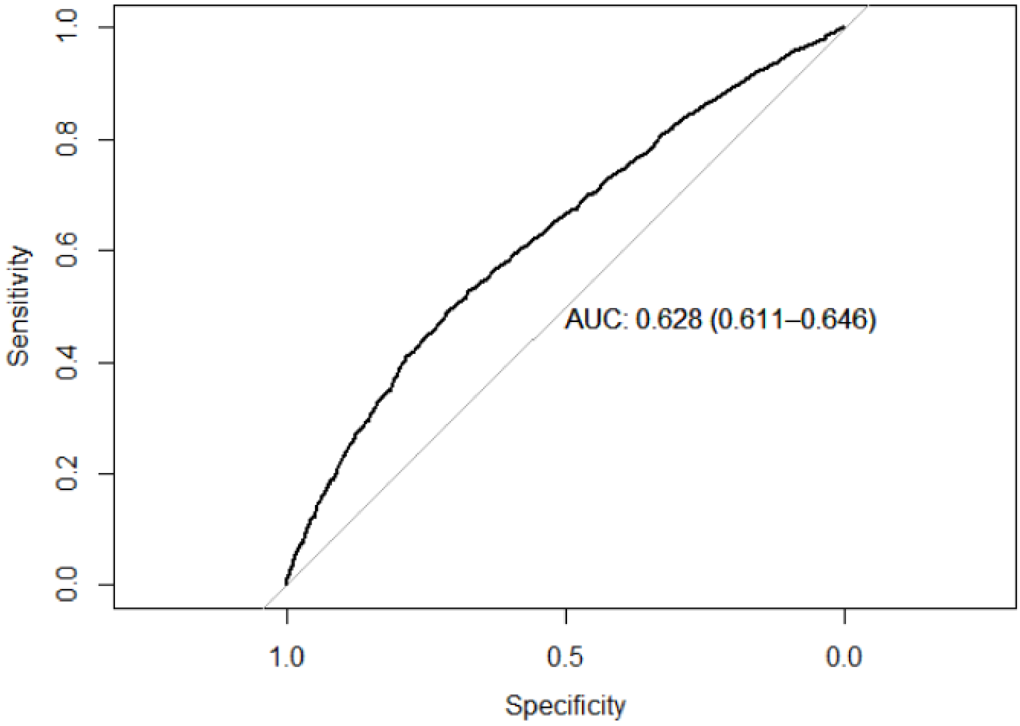

b)

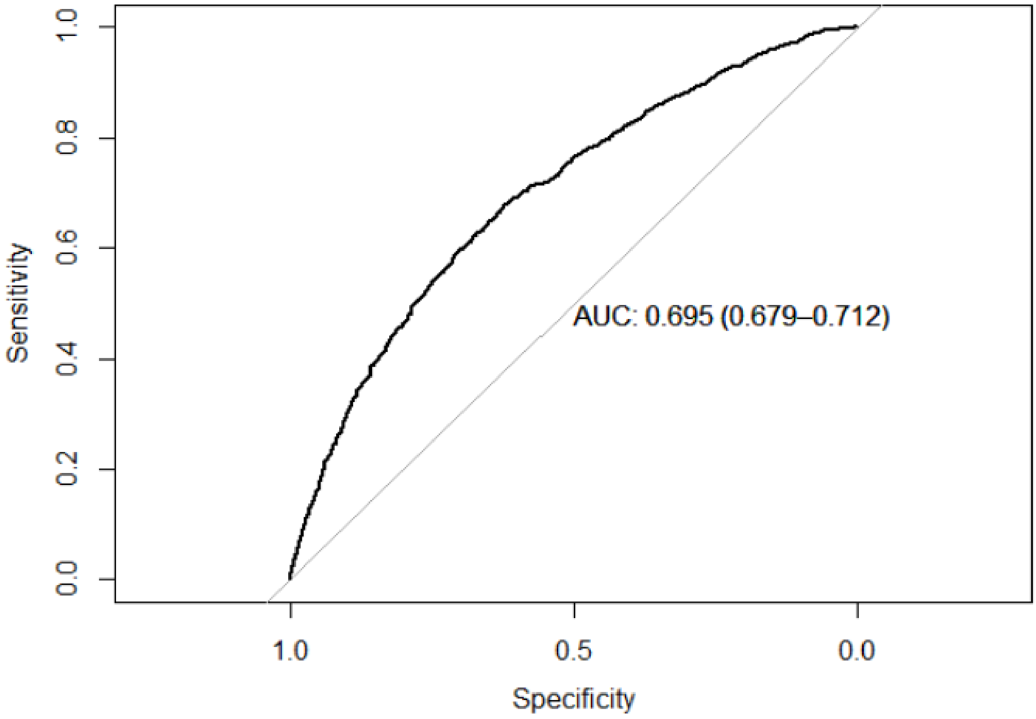

**Figure S3.** A) ROC curve for LOY for age, smoking and alcohol use. B) ROC curve for LOY for age, smoking alcohol use and PRS.

**Figure S4.**

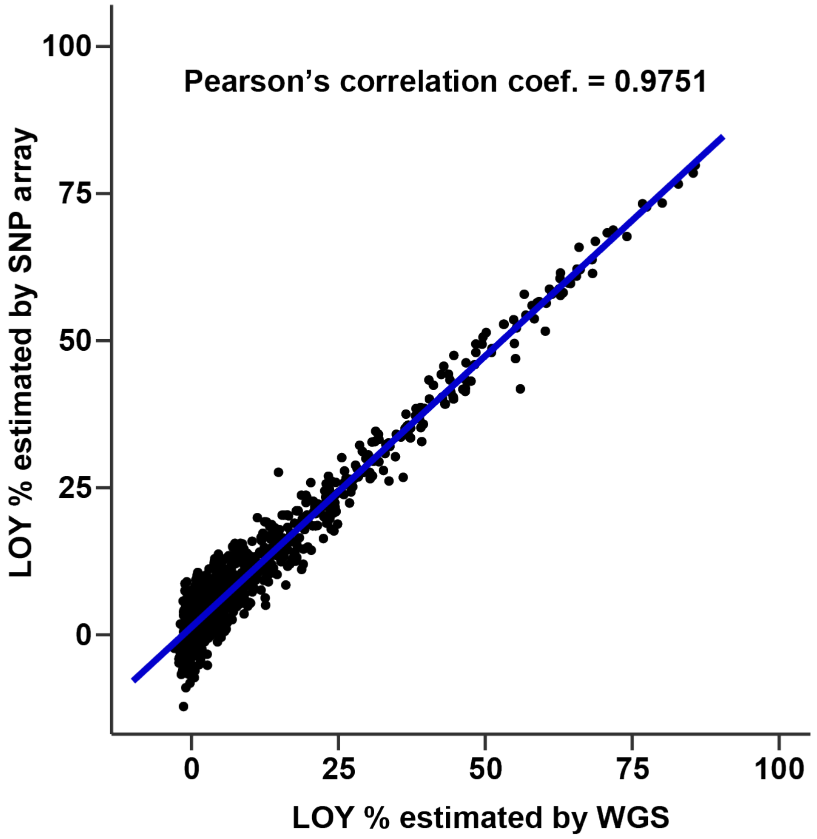

**Figure S4.** Correlation plot comparing microarray-derived LOY calls (Y axis) versus WGS-derived LOY calls (X-axis).

**Figure S5.**

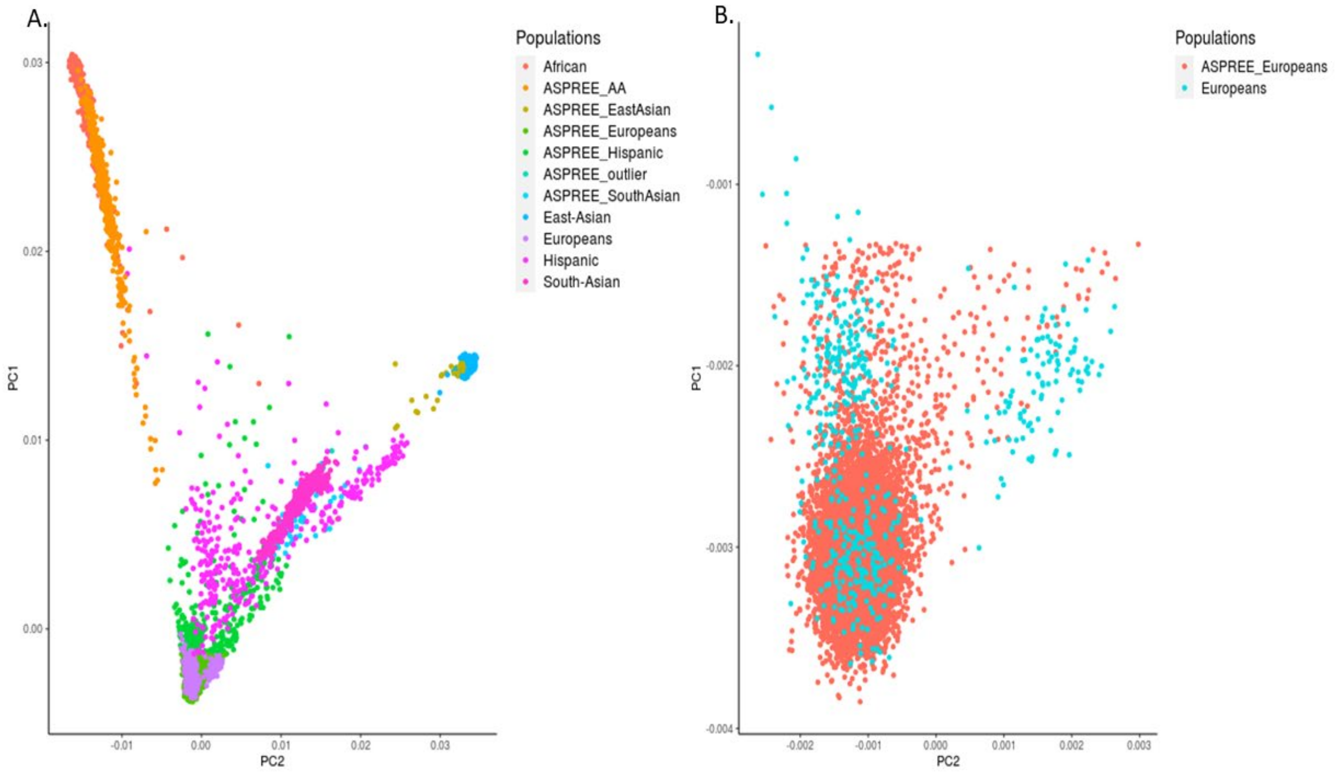

**Figure S5.** Principal component analysis (PCA) of the ASPREE cohort compared with the 1000 Genomes Project. A) PCA plot of all ASPREE participants projected onto 1000 Genome populations. B). PCA plot of ASPREE Europeans samples projected onto 1000 Genome Europeans samples that were included in this study. ASPREE_AA represents African American samples.

**Figure S6.**

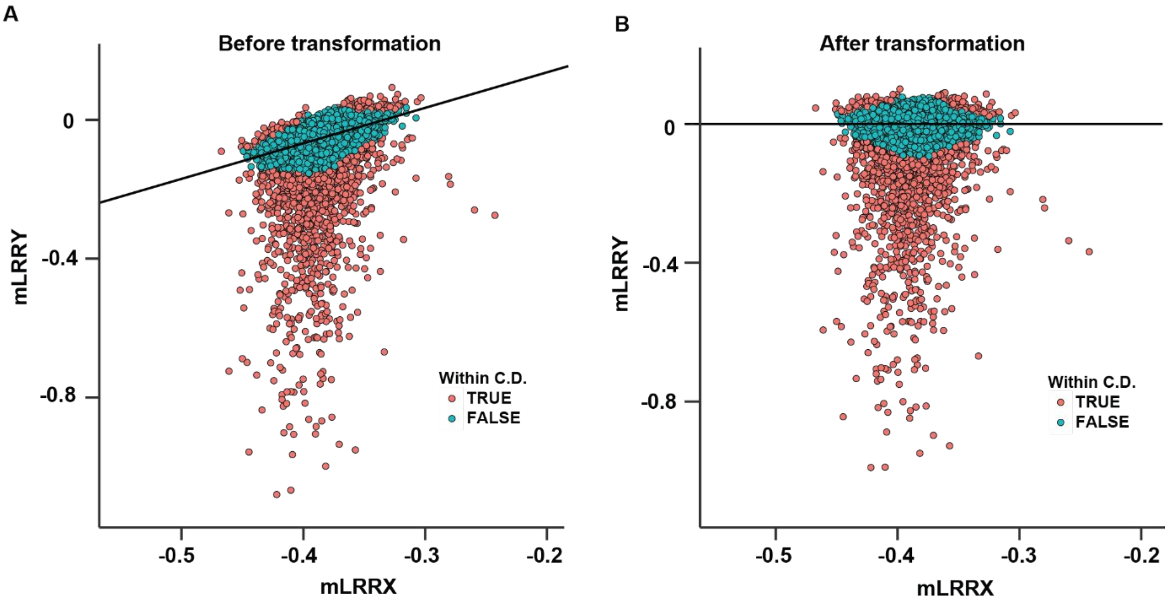

**Figure S6.** Transformation of mLRRY values by adjusting for technical variation as estimated by mLRRX, panels A and B show the unadjusted adjusted datasets, respectively. The correction was based on the coefficients of a linear model estimated between mLRRY and mLRRX in the unadjusted dataset (panel A). To optimize the model only samples showing a value of mLRRY > -0.2 in the unadjusted dataset and samples within a Cook's distance (i.e. similarity of mLRRY and mLRRX estimates between samples, abbreviated C.D.) less than 1.5 IQR of the third quartile, were included in the model. The coefficients of the produced linear regression model could thereafter be used to adjust the mLRRY values [mLRRY_adj = (mLRRY_unadj – 1.023611 · mLRRX) - 0.341685].

**Figure S7.**

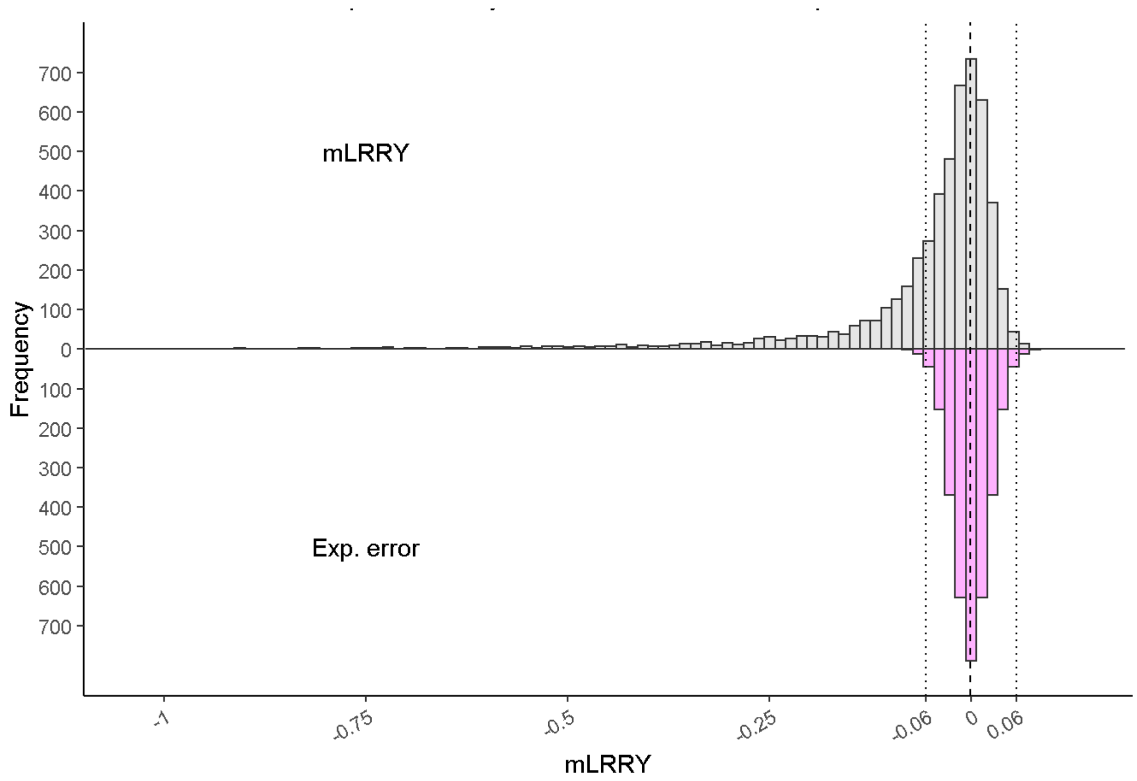

**Figure S7.** Distribution and estimation of mosaic loss of chromosome in ASPREE study from SNP array data. The grey bars show the distribution of mLRRY values observed in the 5.131 investigated samples. The pink bars represents the part of the total variation originating from technical factors, generated by imposing the observed variation in the positive part of the grey tail into a reflected negative tail, while assuming that this variation was distributed symmetrically. The dotted lines marks the 99% confidence limits of the latter distribution and samples were defined as having LOY when the mLRRY value was below the lower 99% confidence limit (-0.06) of this error distribution.

**Figure S8.**

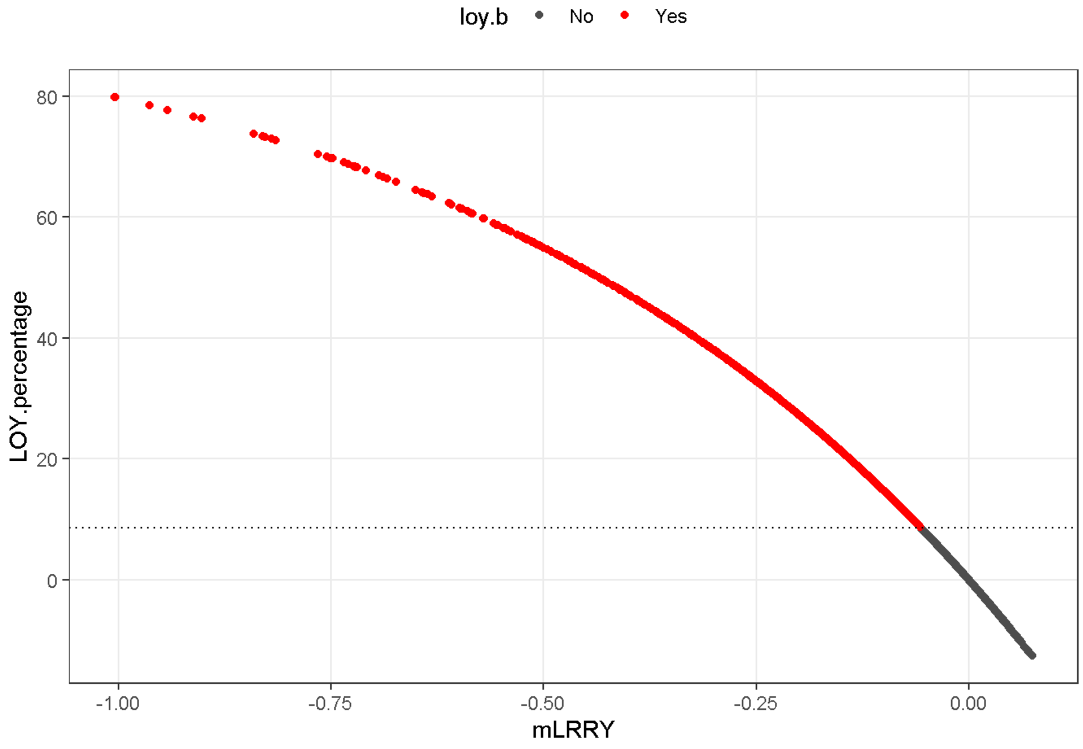

**Figure S8.** Transformation of mLRRY to percentage of cells with LOY. The mLRRY values were converted to LOY percentage using the formula [LOY(%) = $100\cdot(1-2^{2.3\cdot mLRRY})$]. This is a modified version of the published formula [LOY(%) = $100\cdot(1-2^{mLRRY})$] that was optimized from data generated by Illumina arrays. The mLRRY to LOY percentage transformation was derived empirically by optimizing alpha and beta in [(LOY percentage from WGS) = $100\cdot(1-\text{alpha}^{\text{beta}\cdot mLRRY})$] using the data in the present analysis. In this model, alpha and beta was originally estimated as 1.006543 and 2.325276, respectively. To avoid overfitting, both values were rounded to produce the modified formula used here. Samples scored with LOY in at least 8.57% blood cells without chromosome Y (i.e. mLRRY>-0.06) are plotted with red dots.

**Table S1.** Genome wide significant SNPs used to generate the LOY-PRS in ASPREE.

| Consensus Gene | SNP | CHR | BP_hg38 | Risk Allele | OR [Confidence Intervals] |
|---|---|---|---|---|---|
| ARHGAP25 | rs10048745 | 2 | 68735005 | A | 1.1 [1.08-1.12] |
| CTSK | rs10305667 | 1 | 150859632 | C | 1.05 [1.04-1.07] |
| CCND3 / GUCA1B | rs10456506 | 6 | 42048508 | T | 1.14 [1.12-1.17] |
| CCND2 | rs1049612 | 12 | 4303596 | A | 1.05 [1.04-1.07] |
| PMF1 | rs1052053 | 1 | 156232382 | G | 1.16 [1.14-1.18] |
| SESN3 | rs10831321 | 11 | 95231179 | G | 1.05 [1.03-1.06] |
| LTBR | rs10849448 | 12 | 6384185 | G | 1.09 [1.07-1.1] |
| PARP11 | rs11062924 | 12 | 3933197 | T | 1.08 [1.06-1.1] |
| SETBP1 | rs11082396 | 18 | 44500755 | C | 1.23 [1.2-1.25] |
| KRBA1 | rs111725880 | 7 | 149706135 | C | 1.15 [1.12-1.17] |
| MDM2 / NUP107 | rs11177383 | 12 | 68815509 | C | 1.05 [1.04-1.07] |
| BEND7 | rs11258419 | 10 | 13494086 | G | 1.04 [1.03-1.06] |
| HDAC7 / VDR | rs113736796 | 12 | 47819937 | G | 1.12 [1.08-1.16] |
| HEATR3 | rs11642909 | 16 | 50022787 | A | 1.05 [1.03-1.07] |
| SPDL1 | rs116483731 | 5 | 169588475 | G | 1.42 [1.31-1.53] |
| CXCR4 | rs11679328 | 2 | 136126891 | T | 1.15 [1.11-1.18] |
| FLT3 | rs117145034 | 13 | 28100660 | A | 1.25 [1.18-1.33] |
| IKZF1 | rs11769630 | 7 | 50218107 | A | 1.14 [1.11-1.17] |
| FAM117A / SPOP | rs118035610 | 17 | 49726945 | C | 1.26 [1.22-1.3] |
| MRPS18A / VEGFA / MAD2L1BP | rs11965885 | 6 | 43725357 | G | 1.05 [1.03-1.06] |
| SLC25A37 | rs12549737 | 8 | 23536948 | T | 1.07 [1.05-1.09] |
| ZBTB20 | rs12695310 | 3 | 114855180 | G | 1.07 [1.06-1.09] |
| SENP7 / PCNP | rs13062095 | 3 | 101548541 | C | 1.12 [1.1-1.13] |
| SETD2 / NBEAL2 | rs13063578 | 3 | 47046347 | A | 1.06 [1.05-1.08] |
| ANAPC5 | rs13141 | 12 | 121318281 | G | 1.25 [1.17-1.34] |
| HABP4 | rs13286011 | 9 | 96492112 | C | 1.13 [1.08-1.17] |
| SETBP1 | rs141777833 | 18 | 44295910 | C | 1.39 [1.28-1.51] |
| TSPAN9 | rs147764594 | 12 | 3125211 | AT | 1.12 [1.09-1.14] |
| ITPR2 | rs149752564 | 12 | 26435050 | A | 1.12 [1.09-1.15] |
| DAP | rs1531842 | 5 | 10675654 | T | 1.05 [1.03-1.07] |
| PRDM16 | rs1569419 | 1 | 3080038 | T | 1.06 [1.05-1.08] |
| FAM49A | rs16982394 | 2 | 16443481 | A | 1.11 [1.1-1.13] |
| AHI1 | rs17064495 | 6 | 135419391 | C | 1.15 [1.1-1.2] |
| RPN1 / GATA2 | rs17255991 | 3 | 128637371 | C | 1.12 [1.09-1.15] |
| BCL2 | rs17758695 | 18 | 63253621 | C | 1.75 [1.67-1.82] |
| BAX | rs1805419 | 19 | 48955847 | G | 1.07 [1.05-1.09] |
| SETBP1 | rs1849209 | 18 | 44581678 | T | 1.19 [1.17-1.21] |
| CHEK2 | rs186430430 | 22 | 28707610 | C | 2.02 [1.72-2.38] |
| CTBP2 | rs1926785 | 10 | 125161466 | A | 1.07 [1.05-1.09] |
| TET2 | rs199741557 | 4 | 104943372 | ACT | 1.13 [1.09-1.18] |
| KIT | rs218264 | 4 | 54542708 | A | 1.06 [1.04-1.07] |
| MAD1L1 | rs2280548 | 7 | 1937031 | T | 1.13 [1.11-1.14] |

| | | | | | |
|---|---|---|---|---|---|
| MADD / DDB2 | rs2291119 | 11 | 47276650 | C | 1.05 [1.03-1.07] |
| MECOM | rs2293661 | 3 | 169120620 | G | 1.07 [1.06-1.09] |
| NFKB1 | rs230533 | 4 | 102528926 | G | 1.05 [1.04-1.07] |
| MBD5 | rs2382230 | 2 | 148545695 | T | 1.05 [1.03-1.06] |
| CCDC26 | rs2395902 | 8 | 129599841 | G | 1.06 [1.04-1.07] |
| PHF11 / RCBTB1 | rs2407797 | 13 | 49684885 | A | 1.06 [1.05-1.08] |
| AZGP1 | rs2527884 | 7 | 99965219 | G | 1.05 [1.03-1.06] |
| PRDM16 | rs2651932 | 1 | 3182097 | G | 1.08 [1.05-1.11] |
| SGMS1 | rs2688887 | 10 | 50302463 | T | 1.05 [1.03-1.06] |
| TRPT1 | rs2701539 | 11 | 64195906 | G | 1.09 [1.06-1.13] |
| SETBP1 | rs2852780 | 18 | 44697023 | A | 1.08 [1.06-1.1] |
| TCL1A | rs2887399 | 14 | 95714358 | G | 1.25 [1.22-1.27] |
| MAD2L1 | rs2908986 | 4 | 120063721 | A | 1.05 [1.03-1.06] |
| CDK5RAP1 | rs291699 | 20 | 33394393 | T | 1.12 [1.09-1.14] |
| RBPMS | rs2979469 | 8 | 30427575 | C | 1.09 [1.08-1.11] |
| TAL1 | rs34087210 | 1 | 47224170 | G | 1.05 [1.03-1.06] |
| TESC | rs747692613 merged into rs34344826 | 12 | 117040962 | TTCC | 1.05 [1.03-1.07] |
| TP53INP1 | rs34624977 | 8 | 94954676 | CT | 1.06 [1.05-1.08] |
| RPN1 / GATA2 | rs34890930 | 3 | 128610380 | T | 1.06 [1.04-1.07] |
| NFE2 / HNRNPA1 | rs35979828 | 12 | 54292096 | T | 1.14 [1.11-1.17] |
| HMHA1 | rs36084354 | 19 | 1079960 | A | 1.11 [1.08-1.14] |
| CENPN | rs3743503 | 16 | 81022836 | G | 1.14 [1.11-1.16] |
| H2AFY | rs3756364 | 5 | 135386511 | G | 1.08 [1.06-1.11] |
| BCL2L11 | rs3761704 | 2 | 111146121 | G | 1.07 [1.05-1.09] |
| TRIM58 | rs3811444 | 1 | 247876149 | C | 1.06 [1.04-1.08] |
| QKI | rs381500 | 6 | 164057356 | C | 1.16 [1.14-1.18] |
| MYB | rs397687328 | 6 | 135201210 | AT | 1.06 [1.05-1.08] |
| TSC22D2 | rs59633341 merged into rs397689554 | 3 | 150301093 | A | 1.24 [1.22-1.27] |
| CEBPA | rs79966604 merged into rs398101182 | 19 | 33261752 | G | 1.07 [1.05-1.09] |
| USP28 / HTR3A | rs4288784 | 11 | 113874886 | G | 1.09 [1.06-1.12] |
| CDKN1C | rs458069 | 11 | 2837570 | G | 1.05 [1.03-1.06] |
| GACAT3 | rs4669037 | 2 | 16088807 | T | 1.06 [1.05-1.08] |
| SPRED2 | rs4671127 | 2 | 65335836 | C | 1.05 [1.03-1.06] |
| CCND3 / GUCA1B | rs4714550 | 6 | 42018535 | C | 1.1 [1.08-1.12] |
| PHGDH / NOTCH2 | rs478093 | 1 | 119712503 | G | 1.05 [1.03-1.06] |
| PSMA6 / NFKBIA | rs4981287 | 14 | 35358992 | A | 1.08 [1.06-1.1] |
| USP35 / GAB2 | rs760622583 merged into rs55765495 | 11 | 78213610 | T | 1.07 [1.05-1.09] |
| NREP | rs56116444 | 5 | 111726150 | G | 1.18 [1.15-1.21] |
| TERT | rs56345976 | 5 | 1276758 | G | 1.04 [1.03-1.06] |
| KCNA3 / RP11-284N8.3 | rs56795609 | 1 | 110666096 | A | 1.06 [1.04-1.08] |
| HMGA1 | rs57026767 | 6 | 34251921 | C | 1.06 [1.04-1.08] |
| SNTA1 | rs575653337 | 20 | 33419196 | T | 1.24 [1.17-1.31] |
| SYNGR1 | rs5757613 | 22 | 39319257 | A | 1.05 [1.03-1.07] |
| CEBPB | rs6020413 | 20 | 50279908 | T | 1.05 [1.03-1.07] |

| | | | | | |
|---|---|---|---|---|---|
| PMAIP1 | rs60781426 | 18 | 59893578 | A | 1.07 [1.04-1.09] |
| CDKN1C | rs60808706 | 11 | 2836003 | G | 1.11 [1.07-1.15] |
| HM13 / BCL2L1 | rs6089050 | 20 | 31725142 | C | 1.17 [1.15-1.2] |
| PARP11 | rs609018 | 12 | 3962735 | G | 1.09 [1.08-1.11] |
| TSC1 | rs621940 | 9 | 132994743 | C | 1.09 [1.06-1.11] |
| UQCRC1 | rs62618742 | 3 | 48601368 | C | 1.28 [1.22-1.33] |
| SMC2 | rs6479226 | 9 | 104210591 | T | 1.05 [1.03-1.06] |
| KLHL1 | rs670180 | 13 | 70662479 | A | 1.05 [1.04-1.07] |
| FANCL | rs6731121 | 2 | 58764733 | G | 1.05 [1.04-1.07] |
| JARID2 | rs6924733 | 6 | 15385050 | G | 1.05 [1.03-1.06] |
| ARID1B | rs6939093 | 6 | 156655411 | A | 1.12 [1.08-1.16] |
| KANK1 | rs7035770 | 9 | 668887 | A | 1.04 [1.03-1.06] |
| ESYT2 / NCAPG2 | rs710422 | 7 | 158753852 | G | 1.07 [1.05-1.09] |
| NPAT / ATM | rs7129527 | 11 | 108174268 | G | 1.1 [1.08-1.11] |
| ATP8B4 | rs7172615 | 15 | 50065546 | G | 1.07 [1.05-1.08] |
| DLK1 / MEG3 | rs72698720 | 14 | 100712378 | C | 1.18 [1.15-1.2] |
| HM13 / BCL2L1 | rs7271671 | 20 | 31577985 | C | 1.22 [1.18-1.26] |
| SETBP1 | rs72899729 | 18 | 44461166 | G | 1.45 [1.36-1.55] |
| LY75 | rs72955755 | 2 | 159861821 | G | 1.06 [1.04-1.07] |
| ACVR1B | rs73111522 | 12 | 51910937 | A | 1.15 [1.11-1.19] |
| ELF1 / RGS17P1 | rs73176930 | 13 | 41005673 | A | 1.1 [1.08-1.12] |
| STMP1 | rs73721669 | 7 | 135662359 | C | 1.16 [1.14-1.19] |
| TM6SF2 | rs739846 | 19 | 19308262 | A | 1.09 [1.06-1.12] |
| ZBED4 | rs7410534 | 22 | 49896277 | T | 1.09 [1.06-1.12] |
| UGCG | rs74845559 | 9 | 111896656 | C | 1.09 [1.06-1.11] |
| KDELC2 / ATM | rs74911261 | 11 | 108486410 | A | 1.2 [1.15-1.26] |
| FLT3 | rs76428106 | 13 | 28029870 | C | 1.28 [1.2-1.37] |
| ZWILCH / MAP2K1 | rs76428668 | 15 | 66546944 | G | 1.06 [1.04-1.07] |
| IKZF1 | rs7781977 | 7 | 50306538 | C | 1.07 [1.05-1.08] |
| TP53 | rs78378222 | 17 | 7668434 | G | 1.77 [1.65-1.88] |
| SYT4 | rs80036149 | 18 | 43486613 | A | 1.07 [1.04-1.09] |
| MUC22 | rs9262570 | 6 | 31044708 | T | 1.18 [1.13-1.24] |
| CD164 / CCDC162P | rs9400267 | 6 | 109278961 | T | 1.13 [1.12-1.15] |
| KITLG | rs980381 | 12 | 88360793 | T | 1.06 [1.04-1.08] |
| RPN1 / GATA2 | rs9844706 | 3 | 128661938 | G | 1.11 [1.09-1.14] |
| CCDC102A | rs9939347 | 16 | 57536792 | G | 1.06 [1.05-1.08] |
| PARP1 | rs9943081 | 1 | 226350353 | C | 1.06 [1.04-1.08] |

**Table S2.** Frequency of mosaic LOY in blood leukocytes among 5.131 ASPREE male participants, stratified by age groups.

| | Age Categories | | | |
|---|---|---|---|---|
| | **70-74 years** (N = 3144) | **75-79 years** (N = 1280) | **80-84 years** (N = 551) | **>=85 years** (N = 156) |
| **LOY Yes** | 668 (21.0%) | 403 (31.5%) | 214 (43.7%) | 80 (51.3%) |
| **LOY No** | 2476 (79.0%) | 877 (68.5%) | 310 (56.3%) | 76 (48.7%) |

**Table S3.** Area under the curve for each variable for mLRRY derived LOY risk prediction.

| Variable | *AUC (95% confidence interval ) |
|---|---|
| Age | 0.620 (0.60; 0.63) |
| Smoking | 0.526 (0.51; 0.54) |
| Alcohol use | 0.510 (0.50; 0.52) |
| LOY-PRS | 0.642 (0.62; 0.65) |

*Area under the curve using ROC package in R.

**Table S4.** Association of a polygenic risk score for LOY predisposition (LOY-PRS) as a categorical variable (low, middle, high) with mosaic LOY modelled as a continuous variable.

| | *OR (95% CI) | P value |
|---|---|---|
| **LOY low risk PRS** | **Reference** | |
| **LOY middle risk PRS** | 1.45 (1.27; 1.66) | **<0.0001** |
| **LOY high risk PRS** | 2.90 (2.46; 3.41) | **<0.0001** |
| **Age, (years)** | 1.10 (1.08; 1.11) | **<0.0001** |
| **Smoking** | | |
| Never/Former | Reference | |
| Current | 2.19 1.65; 2.90) | **<0.0001** |
| **Alcohol** | | |
| Never/Former | Reference | |
| Current | 1.22 (1.05; 1. 41) | **0.006** |

*Estimated change in the mean of LOY using linear regression. The PRS risk group define as: low risk PRS <20%, Middle risk 30-60% and high risk is >80% PRS distribution.

### SUPPLEMENTARY REFERENCES

1. Genomes Project, C. *et al.* A global reference for human genetic variation. *Nature* **526**, 68-74 (2015).
2. Purcell, S. *et al.* PLINK: a tool set for whole-genome association and population-based linkage analyses. *Am J Hum Genet* **81**, 559-75 (2007).
3. Zheng, X. *et al.* A high-performance computing toolset for relatedness and principal component analysis of SNP data. *Bioinformatics* **28**, 3326-8 (2012).
4. Cooper, N.J. *et al.* Detection and correction of artefacts in estimation of rare copy number variants and analysis of rare deletions in type 1 diabetes. *Hum Mol Genet* **24**, 1774-90 (2015).
5. Forsberg, L.A. *et al.* Mosaic loss of chromosome Y in peripheral blood is associated with shorter survival and higher risk of cancer. *Nat Genet* **46**, 624-8 (2014).
6. Danielsson, M. *et al.* Longitudinal changes in the frequency of mosaic chromosome Y loss in peripheral blood cells of aging men varies profoundly between individuals. *Eur J Hum Genet* **28**, 349-357 (2020).
7. Lacaze, P. *et al.* Medically actionable pathogenic variants in a population of 13,131 healthy elderly individuals. *Genetics in Medicine*, 1-4 (2020).
8. Pinese, M. *et al.* The Medical Genome Reference Bank contains whole genome and phenotype data of 2570 healthy elderly. *Nature communications* **11**, 1-14 (2020).
9. Van der Auwera, G.A. *et al.* From FastQ data to high confidence variant calls: the Genome Analysis Toolkit best practices pipeline. *Curr Protoc Bioinformatics* **43**, 11 10 1-11 10 33 (2013).
10. Boeva, V. *et al.* Control-FREEC: a tool for assessing copy number and allelic content using next-generation sequencing data. *Bioinformatics* **28**, 423-425 (2012).
